# Post operative acute kidney injury in abdominal Surgeries – A retrospective analysis of single center in western India

**DOI:** 10.1101/2021.01.05.21249252

**Authors:** Bhavin Vasavada, Hardik Patel

## Abstract

**AIM:** The aim of our study was to evaluate the incidence and causative factors for acute kidney injury in abdominal surgeries.

**Material and Methods:** All the abdominal surgeries performed between April 2018 to December 2020, in our institution have been analyzed for acute kidney injury. Acute kidney injury defined according to acute kidney injury network classification. Categorical variables were evaluated by chi-square t-test or fisher’s t-test wherever appropriate and continuous variables by Mann Whitney U test for nonparametric data and student t-test for parametric test after skewness and kurtosis analysis. Statistical analysis was done using SPSS version 23. P< 0.05 was considered statistically significant.

**Results:** We performed 402 gastrointestinal and hepatobiliary surgery from April 2018 to December 2020. After exclusion 372 patients were included in the study population. 20 patients (5.37%) were defined as having acute kidney injury according to acute kidney injury network classifications. On univariate analysis acute kidney injury was associated with open surgery (p= 0.003), Intraoperative hypotension (p<0.001), Colorectal surgeries (p<0.0001), Emergency surgery (p=0.028), CDC grade of surgery (p<0.001), increased used to blood products (p=0.001), higher ASA grade (p<0.0001), increased operative time(p<0.0001). On multivariate logistic regression analysis higher ASA grade (p<0.0001) and increased operative time (0.049) independently predicted acute kidney injury. Acute kidney injury was also significantly associated with 90 days mortality. (p= <0.0001).

**Conclusion:** Post-operative acute kidney injury was associated with significant mortality in abdominal surgery. Higher ASA grades and increased operative time predicted acute kidney injury.

## Background

Post Operative Acute kidney injury is one of the very common post-operative complications [1]. Many studies have shown that postoperative acute kidney injury is associated with other morbidities and mortalities. [2,3,4,5].

Very few studies have evaluated the risk of acute kidney injury in abdominal surgeries in the Indian population.

### Aim of the study

The aim of this study was to evaluate the incidence and risk factors of acute kidney injury retrospectively from our data of abdominal surgeries.

## Material and Methods

### Inclusion criteria

All patient who underwent gastrointestinal and hepatobiliary surgeries.

### Exclusion criteria

· Patients who had acute kidney injury preoperatively
· Patients who were on dialysis before surgery
· Patients who died on postoperative day 1, before Acute Kidney Injury criteria are fulfilled.

All the gastrointestinal surgeries performed from April 2018 to December 2020 in our institution have been analyzed for acute kidney injury.

### Acute kidney injury definition

Acute kidney injury defined according to acute kidney injury network classification. [6,7]. Any grade of acute kidney injury was considered significant. Preoperative and Intraoperative factors were analyzed for the development of acute kidney injury.

### Statistical analysis

Analysis of means or medians were selected according to skewness and standard error of skewness and kurtosis and standard error of kurtosis analysis. Categorical variants were analyzed using the chi-square test or fisher t-test where ever appropriate. Continuous variables were analyzed using the Mann-Whitney U test.

A p-value of less than 0.05 was considered significant. Multivariate analysis was done using a logistic regression method. SPSS (IBM) version 23 was used for statistical analysis.

We also prepared the Kaplan Meier survival curve with log-rank analysis to compare 90 days survival rates in patients with or without AKI.

## Results

We performed 402 gastrointestinal and hepatobiliary surgery from April 2018 to December 2020. After applying exclusion criteria 372 patients were included in the study. 26 patients had preoperative acute kidney injury, 4 patients were on dialysis and one patient died on postoperative day 0 and hence excluded from the study. Various type of abdominal surgeries performed is described in table 1.

**Table 1:** Types of Abdominal surgeries.

| TYPE OF SURGERIES | NUMBERS |
| --- | --- |
| UPPER GI SURGERIES | 17 |
| SMALL BOWEL SURGERIES | 47 |
| HPB SURGERIES | 212 |
| COLORECTAL SURGERIES | 57 |
| HERNIA | 39 |

20 patients (5.3%) were defined as having acute kidney injury according to acute kidney injury network classifications. [6,7]. [Figure 1]

**Figure 1.**
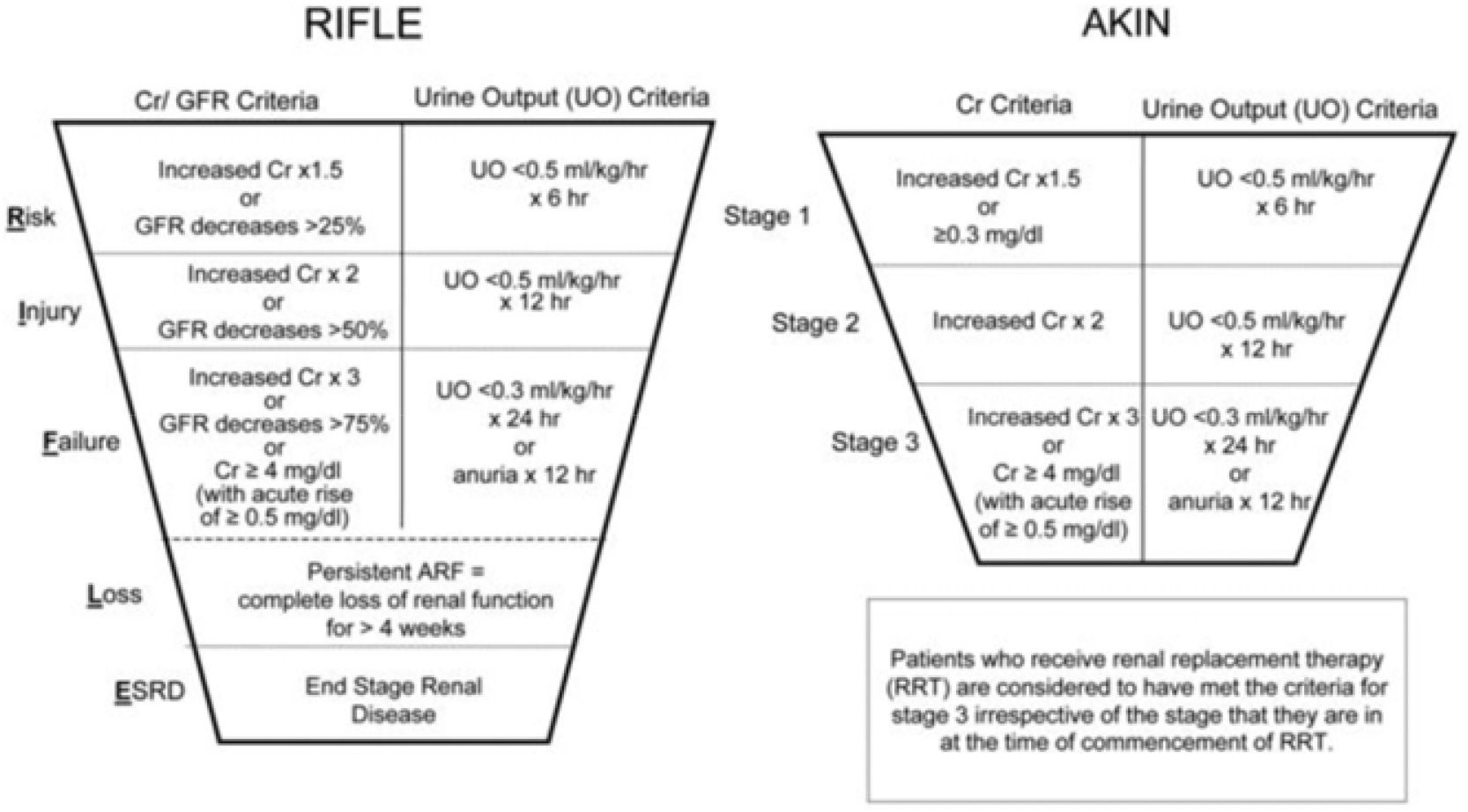
RIFLE and AKIN classifications for acute kidney injury. Risk–Injury– Failure–Loss–End stage renal disease (RIFLE) and Acute Kidney Injury Network (AKIN) classifications for acute kidney injury (adapted from [6,10]).

On univariate analysis acute kidney injury was associated with open surgery (p= 0.003), Intra operative hypotension (p < 0.001), CDC grade of surgery (p<0.0001) [8], Colorectal Surgeries (p< 0.001), emergency surgeries (p=0.028), increased used of blood products (p=0.001),higher ASA grade (p<0.0001), increased operative time (p<0.0001). [Table 2]

**TABLE 2:** UNIVARIATE ANALYSIS OF ACUTE KIDNEY INJURY.

| FACTORS | ACUTE KIDNEY<br>INJURY (AKI)<br>(N=20) | NO ACUTE<br>KIDNEY INJURY<br>(N=352) | P VALUE |
| --- | --- | --- | --- |
| AGE<br>(Median/range) | 58/(34-80) | 54/(7-84) | 0.122 |
| SEX (M/F)<br>(211/161) | (12/8) | 199/153 | 0.760 |
| Malignancy (73) | 6 | 67 | 0.247 |
| Intra Operative<br>hypotension (n=24) | 7 | 17 | <0.001 |
| Emergency<br>Surgeries (n=67) | 7 | 60 | 0.028 |
| HPB surgeries<br>(214) | 13 | 201 | 0.643 |
| Colorectal (56) | 4 | 52 | <0.0001 |
| Small bowel (19) | 3 | 16 | 0.715 |
| Upper GI surgery | 0 | 19 | 1 |
| Hernia | 0 | 37 | 0.239 |
| OPERATIVE<br>TIME<br>(Median/range) | 180 (60-600) | 90 (15-800) | <0.0001 |
| ASA (Mean/range) | 4 (1-4) | 2.00/(1-4) | <0.0001 |
| BLOOD<br>PRODUCT<br>(Median/range) | 1.0/(0-15) | 0.00/(0-40) | 0.001 |
| GRADE OF<br>SURGERY<br>(median/range) | 3.0/(2-4) | 2.0/(1-4) | <0.0001 |
| Open Surgeries.<br>(n= 206) | 16 | 190 | 0.003 |

TABLE: 2 UNIVARIATE ANALYSIS OF ACUTE KIDNEY INJURY.

On multivariate logistic regression analysis higher ASA grade (p=0.001, odds ratio 15.05, 95% confidence interval 4.05-56.20) and increased operative time (p=0.049, odds ratio 1.005, 95% confidence interval 1.001-1.015) independently predicted acute kidney injury. [Table 3]

**TABLE 3:** MULTIVARIATE ANALYSIS OF ACUTE KIDNEY INJURY

| Factor | P value | Odds ratio | 95% confidence interval |
| --- | --- | --- | --- |
| Blood products | 0.223 | 0.828 | 0.61-1.12 |
| ASA | 0.0001 | 15.10 | 4.05-56.2 |
| Operative time | 0.049 | 1.005 | 1.000-1.011 |
| Open Surgery | 0.697 | 1.50 | 0.194-11.63 |
| Grade of surgery | 0.582 | 0.698 | 0.194- 2.51 |
| Intra operative Hypotension | 0.195 | 2.83 | 0.585-13.72 |
| Emergency Surgery | 0.358 | 0.433 | 0.078-2.58 |
| Colorectal surgeries | 0.680 | 1.37 | 0.301-6.298 |

Acute kidney injury was also significantly associated with 90 days mortality. (p= <0.0001)

On Kaplan Meier, survival analysis patients with acute kidney injury had significantly less 90 days survival than patients without acute kidney injury. [Figure 2].

**Figure 2.**
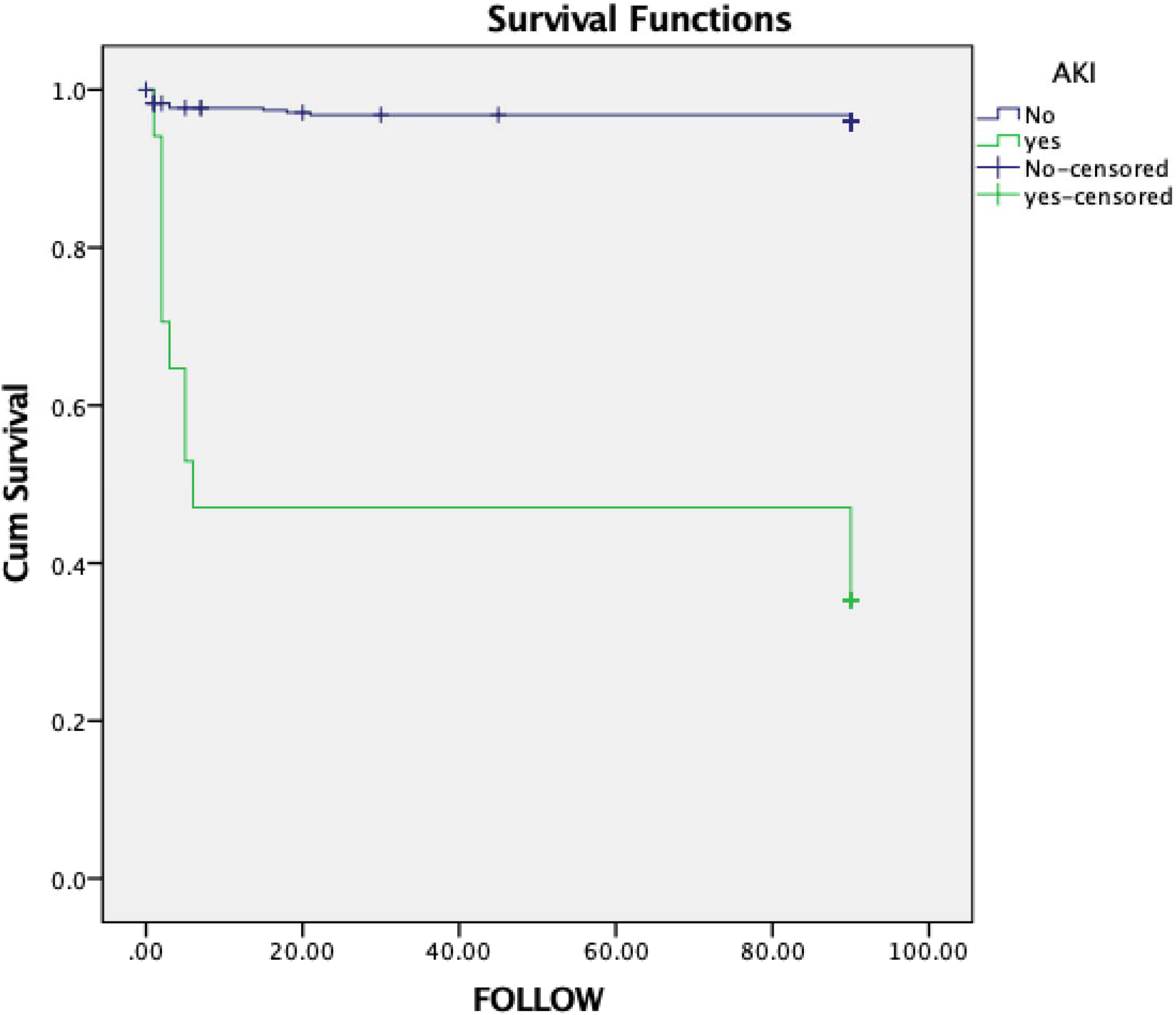
Kaplan meier survival analysis showed 90 days survival was sginificantly less in patient who developed acute kidney injury. (p < 0.0001, log rank analysis)

## DISCUSSION

Acute kidney injury (AKI) is a common complication in patients undergoing major Gastrointestinal and hepatobiliary surgery. Various studies stated the incidence of AKI after major abdominal surgery may go up to 35%. Several patient-related, procedure-related factors for AKI. AKI was associated with increased incidence of morbidity and mortality in various studies. [8,9]

The aim of our study was to evaluate the incidence and various factors associated with Acute kidney injury. Two criteria for diagnosing acute kidney injury are widely used one is RIFLE criteria and the other is Acute Kidney Injury Network criteria. [6,10]. We used Acute Kidney Injury Network criteria to diagnose acute kidney injury network and we included all the grades of acute kidney injury network classification in our analysis. [Figure 2]

The overall incidence of acute kidney injury in our cohort was 5.3%. Cho et al. reported 7.6 percent acute kidney injury after HPB surgeries. [9] Meersch et al. reported a high incidence of acute kidney injury.

Acute kidney injury in our study was significantly associated with mortality. Literature also shows that acute kidney injury is associated with high mortality. [9,10,11,12,13].

On univariate analysis in our study open surgeries, intraoperative hypotension, higher ASA (American Society of anesthesiology) grade, higher CDC wound grade, increase blood product requirement, colorectal surgeries, emergency surgeries, and increased operative time were associated with the development of acute kidney injury. Hobson et al. studied various risk factors associated with perioperative risk factors and showed a previous history of chronic kidney disease and hemoglobin level was associated with postoperative acute kidney injury. [14] In our study increase, blood product required was associated with acute kidney injury. Lim et al. suggested that major hepatectomy, increase meld score, advanced age, and prolonged operative duration were associated with acute kidney injury.[15]

Various studies showed that intraoperative hypotension was associated with post-operative acute kidney injury.[16,17,18] In our study also intraoperative hypotension was associated with acute kidney injury in univariate analysis however it did not independently predict acute kidney injury on multivariate analysis.

In the multivariate analysis in our study higher American society of anesthesiology score (p< 0.001) and increased operative time (p=0.015) were associated with acute kidney injury Literature also supports these findings. [19,20]

In our study, acute kidney injury was associated with increased hospital stay which is also supported by the literature. [21,22]

Being a retrospective analysis, our study has inherent limitations associated with retrospective studies. The number of patients who developed acute kidney injury was low in our studies and that may be the cause of the wide confidence interval of odds ratios after multivariate analysis. The effect size of operative time in predicting acute kidney injury was very limited and that is why it was only marginally significant, higher sample size may change this finding. However, the ASA score was highly significant with a very high odds ratio in predicting acute kidney injury and we believe this finding may not change with increasing sample size. More studies with a higher number of patients are required to confirm our findings. Our earlier data with a limited number of patients (23) also showed Higher ASA scores and increased operative time predicted postoperative acute kidney injury but in that data, operative time was highly significantly associated (p=0.015) with postoperative acute kidney injury but in this updated data it is just borderline significant with p-value of 0.049 and lower limit of 95% confidence interval of odds ratio 1.0. ASA retained its very high significant level with p-value < 0.0001. This shows the importance of sample size and the need for studies with higher sample sizes and adequate power.

In conclusion, postoperative acute kidney injury was associated with significant mortality in gastrointestinal and hepatobiliary surgery. Higher ASA grades and increased operative time independently predicted acute kidney injury in our data.

## Data Availability

Data will provided on demand.

## Notes

### Competing Interest Statement

The authors have declared no competing interest.

### Funding Statement

No funding required.

### Author Declarations

Shalby institutional review board.

